# Chaos theory applied to the outbreak of Covid-19: an ancillary approach to decision-making in pandemic context

**DOI:** 10.1101/2020.04.02.20051441

**Authors:** S. Mangiarotti, M. Peyre, Y. Zhang, M. Huc, F. Roger, Y. Kerr

## Abstract

Predicting the course of an epidemic is difficult, predicting the course of a pandemic from an emerging virus even more so. The validity of most predictive models relies on numerous parameters, involving biological and social characteristics often unknown or highly uncertain. Data of the COVID-19 epidemics in China, Japan, South Korea and Italy were used to build up deterministic models without strong hypothesis. These models were then applied to other countries to identify the closest scenarios in order to foresee their coming behaviour. The models enabled to predict situations that were confirmed little by little, proving that these tools can be efficient and useful for decision-making in a quickly evolving operational context.

## Introduction

The coronavirus SARS-CoV-2 (Severe Acute Respiratory Syndrome Coronavirus 2) is responsible for the Covid-19 epidemic that broke out in Wuhan (China) on December 2019 [1]. Although being identified in the early stages of the epidemic as being close to two other coronaviruses (SARS and MERS) [2], its epidemiological risks in terms of propagation and lethality were not known. Investigations reported by the Chinese Centre for Disease Control and Prevention (China CDC) demonstrated that a new coronavirus was at the origin of this epidemic [3]. The retrospective analysis of the earlier transmission in Wuhan revealed that human-to-human transmission had occurred since the middle of December through close contacts [4] and possibly earlier. After a very rapid dissemination in the Hubei province, the disease has spread to all the other provinces in China. Currently, the epidemic seems to be getting under control thanks to strict control measures [5]. During its early spread in China, the virus also reached several countries in the world, in particular Japan where early measures enabled to control its spread relatively well [6] although restarts are now observed. More recently several important new clusters broke out in South Korea, Iran and Italy by the end of February. Beginning of March most of Europe was affected, to be followed by the USA. Currently the epidemic is affecting the whole world [7].

Various techniques have been developed to model the epidemics of infectious diseases. Most of these are based on compartment models which separate the populations in main classes. The model enables to represent the interactions between these classes based on pre-established mathematical rules. The simplest formulation comes from the early work by Kermack and McKendrick in the 1920s [8] and involves three classes: one for the people sensitive to the disease who are prone to contracting the disease, a second one for the infectious people who have contracted the disease and can infect susceptible people, and a third one for the people outside this cycle either because they became immunised after recovering, or because they left the study area, or finally because they have died.

Although some specific model formulations can be usefully fostered, all the mechanisms may not always be known and therefore the complete formulation of the equations governing an epidemic will generally be unknown. This is especially true when coping with a new disease outbreak. Three main problems will be met in a modelling perspective of such a situation: (1) What are the relevant variables for a given epidemic? (2) What are the governing equations coupling these variables? (3) What are the parameter values of these equations? And, in between these questions, two other very practical questions: (4) what observations do we have to build and constrain a model? And, as a corollary, (5) how to reformulate the governing equations based on the observations we have? (see Suppl. Mat. 1 for a more detailed contextualization)

Based on the chaos theory [9], the global modelling technique [10–13] offers an interesting alternative with respect to other approaches. It is well adapted to the modelling and study of unstable dynamical behaviours: it enables to detect and extract the deterministic component underlying the dynamical behaviour; and, as a consequence, it can be a powerful approach to analyse dynamics which are highly sensitive to the earlier conditions and to detect chaos (see Suppl. Mat. 2). Another interesting aspect of this technique comes from the potential it offers to work even when important variables are missing which is generally the case in epidemiology. Finally, it has proven to be a powerful tool to detect couplings between observed variables, and even, when all the dynamical variables are observed, to retrieve the original algebraic formulation of the governing equations in a compact and potentially interpretable form [14].

Numerous studies have been based on chaos theory to study epidemiological behaviours [15–17] but a global modelling approach *per se* has rarely been applied to biological systems. The first replicable application was in ecology [18]. In epidemiology, it enabled to obtain an interpretable model for the epidemic of Bombay bubonic plague (1896–1911) by extracting the couplings between the human epidemic and the epizootics of two species of rats. Although obtained from observational time series without strong *a priori* model structure, it was found possible in the latter case to give an interpretation to all the model’s terms [19]. A model was also obtained for the West Africa epidemic of Ebola Virus Disease (2013–2016) coupling the two observed variables made available with a regular sampling: the cumulated numbers of infected cases and deaths [20]. In the present study, this modelling approach is used to model the current Covid-19 epidemic in Asia (China, Japan, South Korea) and Italy and then to produce scenarios for fifteen other countries where the disease was introduced later and spread locally. Two general equation forms are considered in the present study: (1) the multivariate form

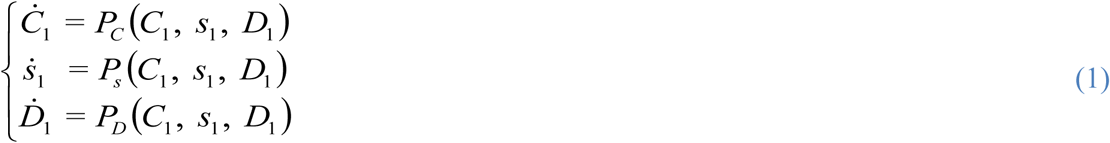

based on three observed variables with the aim to retrieve the couplings between *C*_1_ the number of daily new cases, *s*_1_ the number of daily additional severe cases (positive or negative) and *D*_1_ the number of daily deaths; And (2) the canonical form

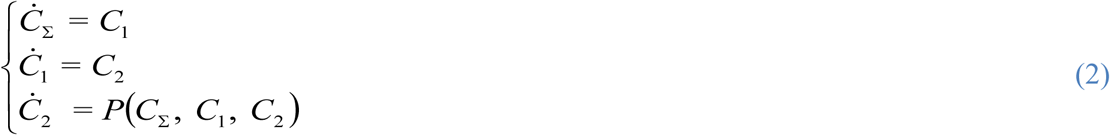

based on a single observed variable – here *C*_Σ_ the cumulative number of new cases since the beginning of the epidemic – and its successive derivatives *C*_1_ and *C*_2_, with the aim to analyse and compare the dynamical behaviours. This latter formulation was also applied to the cumulative number of deaths *D*_Σ_ with its derivatives *D*_1_ and *D*_2_.

Two main data sets were considered for the present study. The official data from the National Health Commission of the People’s Republic of China [21] were used to study the original outbreak at China’s scale (from 21 January to 10 April 2020). The data from the Johns Hudson University [22] were used to monitor the outbreaks at the province’s scale in China and at the country’s scale for other countries. Their description and pre-processing are given with further details in Suppl. Mat. 3 (Fig. S1). Three original time series were considered: (1) the daily cumulated number *C*_Σ_(*t*) of confirmed cases, (2) the daily number *s*(*t*) of severe cases currently under intensive care; and (3) the daily cumulated number *D*_Σ_(*t*) of deaths; from which the derivatives (required for the modelling approach used in the present study) were deduced, corresponding to the daily number of new cases *C*_1_ (*t*), the daily additional severe cases *s*_1_ (*t*) and daily number of new deaths *D*_1_ (*t*) (see Eq. 1).

Considering the set of variables (*C*_1_, *s*_1_, *D*_1_), a model of chaotic behaviour was obtained on February 5 for the window 21 January to 4 February 2020 [23]. Its time evolution is shown in Figure one. This model **M_1_**

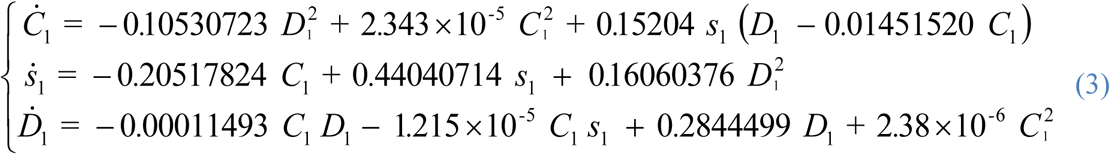

comprises eleven terms. It emphasizes the existence of strong but nonlinear couplings between the three observed variables. Unfortunately, this model is difficult to interpret, very likely because two of the observed variables – the daily numbers of new cases and additional severe cases – can present overlaps, the latter being potentially included in the former one. As a consequence, the present model can only be a reformulation of the original processes as well as a rough reduction.

The numerical integration of the model shows a transient of approximately 15 days that corresponds to the window used to identify the model Fig. 1. Three simulations were performed starting from slightly different initial conditions. These trajectories alternatively come closer and move away one to another but do not converge to a single time evolution which illustrates the high sensitivity to the initial conditions. During the identification period (DoY 20 to 25), the trajectories show a good consistency of the model (grey lines) with the observed data 10 (plain black line). The agreement clearly deteriorates after this period (dashed black line). The data show that the observed number of new cases *C*_1_(*t*) starts decreasing on DoY 37 after a peak of ~5000 cases per day, whereas the simulations continue rising until reaching a steady regime of much larger magnitude (>7000 cases per day). This can be explained as follows. The lockdowns were set on in China from DoY 23 to 29 (Table 1). But considering the incubation period of 3–5 days, the model was thus obtained while the effect of the confinement was still partial, and while the disease was still under progress for contaminated people although contamination had been slowed down. The large oscillations observed in *s*_1_(*t*) (Fig. 1b) are reproduced by the model although slower in the simulations. Finally, although underestimated by the model, the agreement with the data is obviously better for the daily deaths (Fig. 1c). Death is the result of the successive stages of the disease which duration can highly vary from one patient to another. This generates a delay following the peak of new cases. The number of deaths will thus depend on the number of simultaneous cases *s* but also on the health system capacity, in particular on the number of ventilators available in a country. Exceeding the capacity limitation can have a direct effect on deaths generating a plateau rather than a peak for the daily deaths.

**Table 1.**
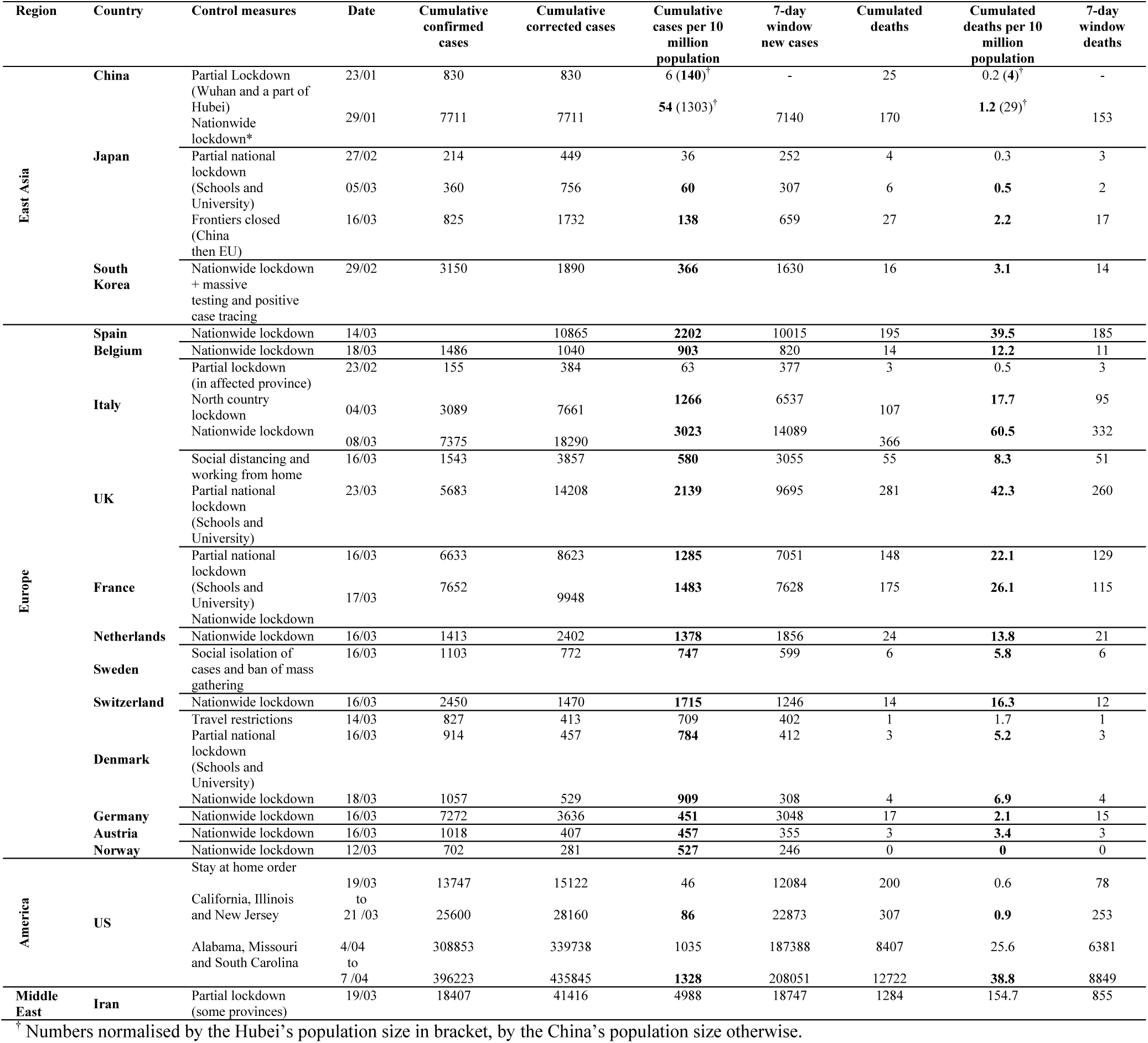
Control measures and epidemic situation. Control measures and epidemic situation Control measures by country, and corresponding dates and epidemic situation in terms of cases and deaths (corrections for country inter-comparisons have been applied, see Suppl. Mat. Data 2).

**Fig. 1.**
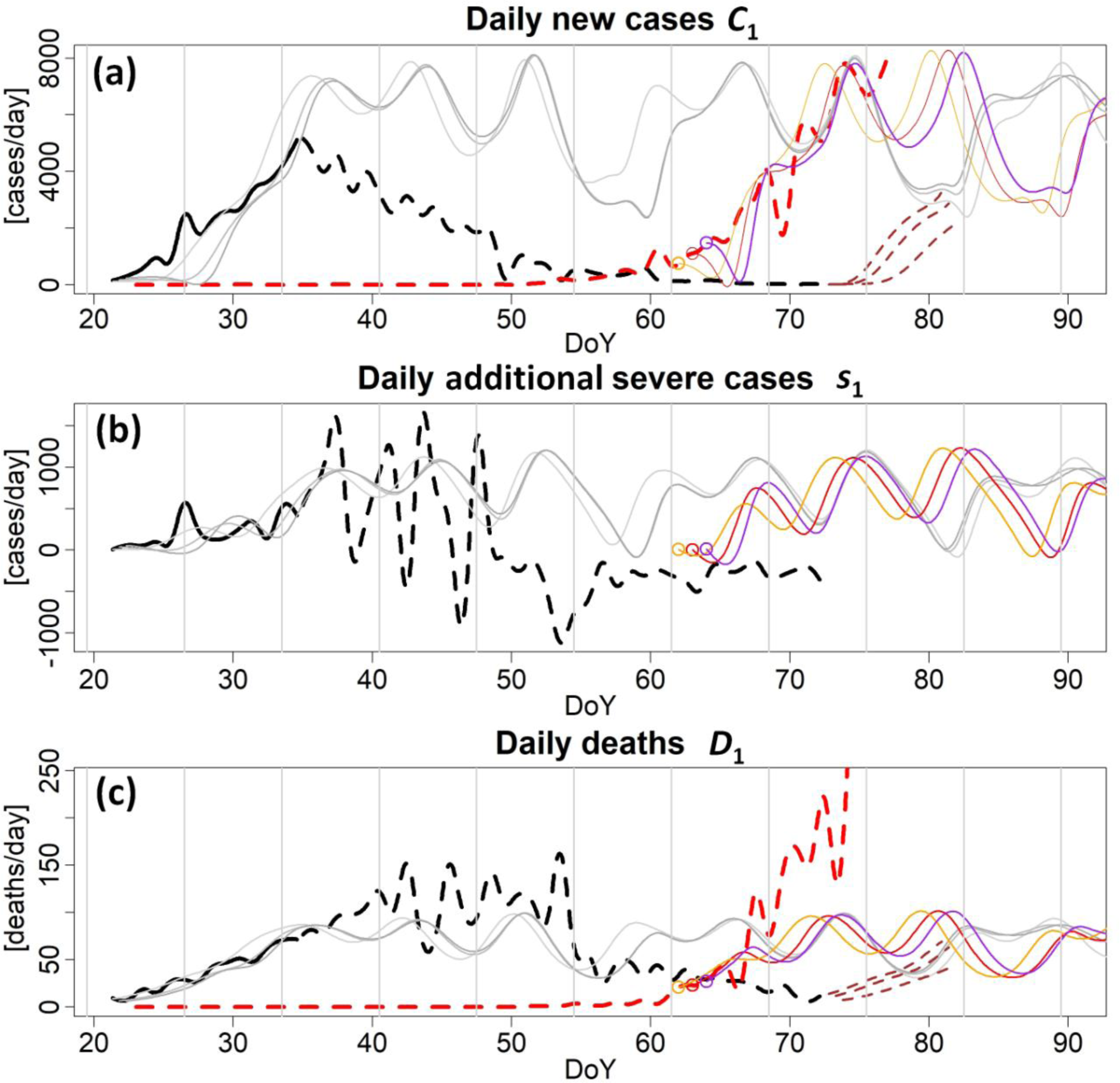
Observed (thick lines) and modelled (light lines) with **M_1_** (Eq. 3) time series for China (black, grey and brown) and Italy (red, orange and purple) due to Covid-19 from 21 January (DoY 21) to 2 April 2020 (DoY 92). The part of the observations used to identify the model is in plain lines. Three variables are presented: the daily number of confirmed new cases *C*_1_ (a), and of additional severe cases variations *s*_1_ (b) and the daily deaths *D*_1_ (c). Note that a correction factor has been applied to the number *C*_1_ of confirmed new cases in Italy to make the comparison with China consistent (see Suppl. Mat. 2). Dashed brown lines correspond to simulations of a possible restart in China.

The representation in the phase space shows that, after the transient, the model trajectory reaches a steady regime characterised by a chaotic attractor (Figure 2). A more detailed analysis of the model (see Suppl. Mat. 4 and Fig. S2) proves that the dynamic of model M_1_ is very close to a phase non-coherent regime, that is, a much less predictable behaviour.

**Fig. 2.**
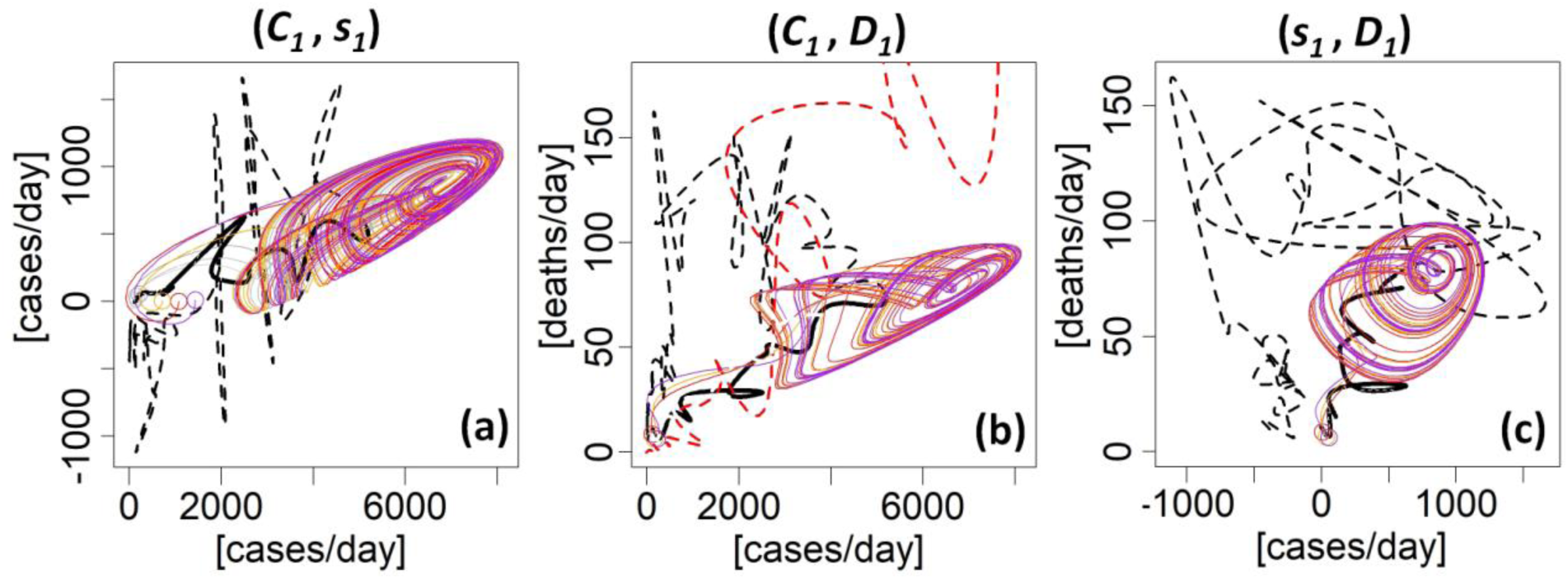
Original and modelled phase portraits. Three projections (*C*_1_, *s*_1_) in (a), (*C*_1_, *D*_1_) in (b) and (*s*_1_, *D*_1_) in (c) of the phase space as reconstructed from the model’s trajectory (colour trajectories). The three colours correspond to different initial conditions (colour circles), each taken from the original data set, on 21 January 7:00 (red), 19:00 (orange) and 22 January 7:00 (purple) 2020. After a 15-day transient, the trajectories converge to a chaotic attractor. Trajectories reconstructed from the observational data are also presented: for all China (in black) and for Italy (in red). The part of the observations used to identify the model is in plain line.

**Fig. 3.**
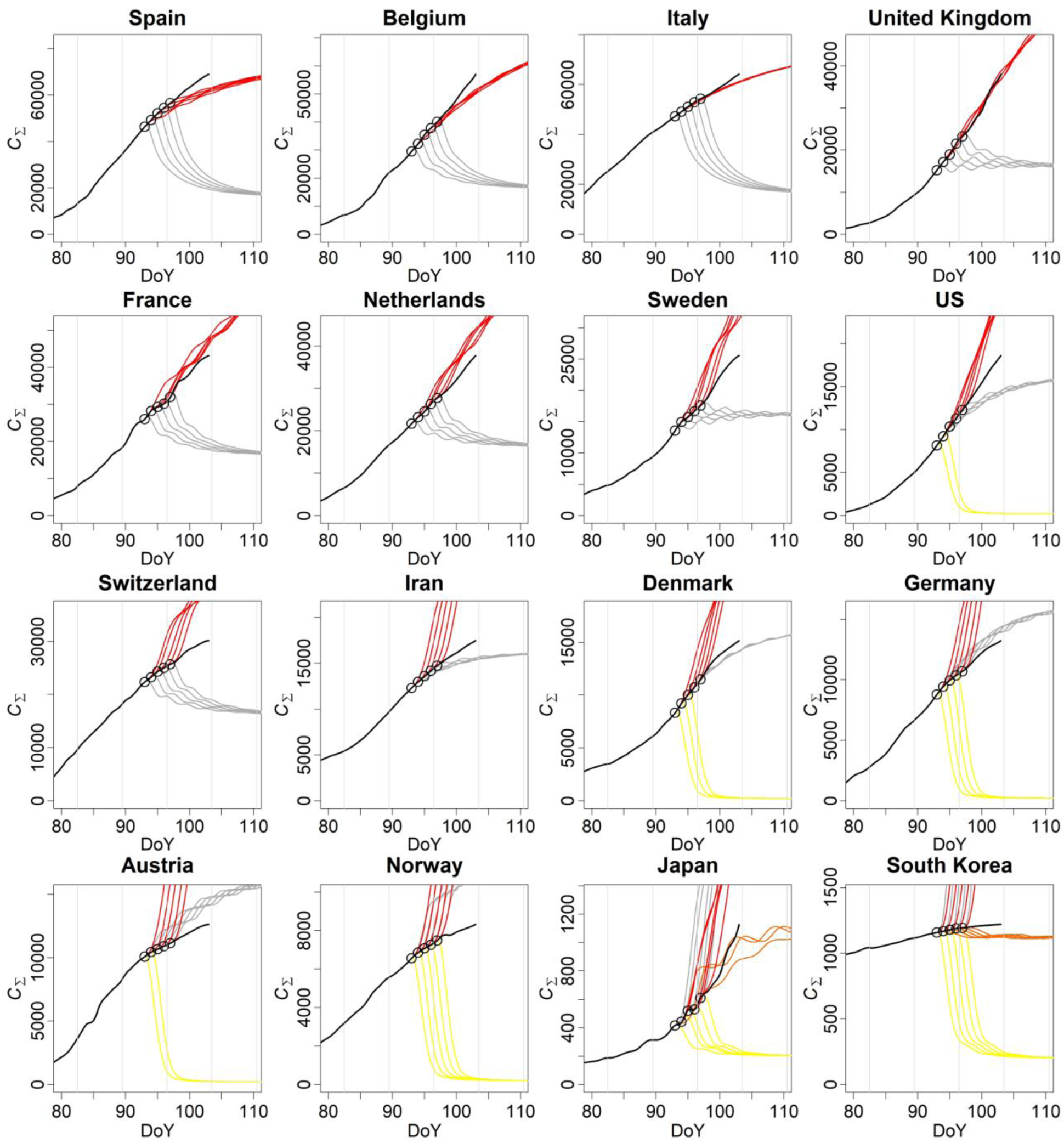
Empirical scenarios simulations (variable *C*_Σ_) Empirical scenarios (in color) of the number of cumulative cases per 10 million population, applied to fifteen countries based on the models obtained for seven Chinese provinces, South Korea, Japan and Italy. Observations are in black plain lines. For each model, an ensemble of five simulations was run starting from the observational initial conditions (black circle) from 2 April (DoY 93) to 6 April (DoY 97). Population size is taken into account but age, geographical distribution and society organization are not. Correction factors were applied to each country to account for the inter-countries discrepancies between the cases and deaths number (see Suppl. Mat. 2). Four main scenarios have been kept: the Jiangxi (in yellow), the South Korea (in orange), the Hubei (in grey) and the Italian scenarios (in red). The other scenarios were rejected automatically.

Four other models **M_2_** to **M_5_** were obtained (see Fig. S3-S6) using a slightly different time window. Interestingly, small parameter changes of model **M_2_** give rise to a restart after a decreasing period (Fig. S4). This illustrates the possibility to withstand the peaks of the outbreak by controlling the proper parameters; It also reveals the capacity of the approach to obtain governing equations enabling a certain diversity of behaviours. Note that all these models (**M_1_** to **M_5_**) could be obtained only for windows at the transition between the exponential propagation of the disease and the beginning of the decrease. Yet, while chaotic models can give rise to unpredictable trajectories, they rely on steady dynamics: their governing equations are invariant. Although lockdowns were applied in China during the period ranging from DoY 23 to 29, due to the incubation time the results of these measures started to become effective around 6 February 2020. It is why these chaotic models could be obtained only on a brief time window, revealing the tipping point from uncontrolled (and unbounded) behaviour to a controlled situation, by the early beginning of February (around DoY 37). And it is also why, soon after (around DoY 41), it became possible to obtain a canonical model (general equation form corresponding to Eq. 2) for the set of variables (*C*_Σ_, *C*_1_, *C*_2_) converging to fixed points (see model **K_1_** in Suppl. Mat. S5, Fig. S7-S8).

At present, despite some restarts, the epidemic seems to be getting under control in China. Since the immunisation remains largely insufficient to modify the dynamic significantly (see Table S2), the obtained models can be used to simulate a restart. Three simulations starting from DoY-72 7pm and DoY-73 7am and 7pm were run (see Figure one in dashed brown lines) showing that a quick restart must be expected if the control measures were to be released at this time.

Using the Italian data as new initial conditions, the model **M_1_** (light coloured lines) was run again to compare it with the evolution observed in Italy (dashed red line) revealing a good agreement during the transient. This agreement may be surprising considering the difference in population size between the two countries (no population normalization was applied here). The behaviour observed in China relies, for more than 83.5%, on the Hubei province which population size is quite similar to that of Italy. Comparing with the evolution of the epidemic in Italy, this simulation is backing up the fact that the strict control measures applied by the Chinese authorities played an important role in the control of the epidemic. Very strict measures were taken in Hubei (total quarantine of Wuhan and of a part of Hubei province) while the number of daily new cases was around 140 per 10 M population in Hubei. Similar measures were taken in Italy (on 8 March 2020) and more recently in Spain (14 March 2020) but comparatively much later, when the number of cases reached (around 3023 per 10 M population in Italy and 2202 in Spain, see Table 1). In France the control measures enforcing confinement of all the French population based on voluntary basis but under government control has arrived much later than in China but slightly earlier than in Italy and Spain in terms of number of cases (~1483).

Using the model **M_1_**, the cumulated counts *C*_Σ_(*t*) and *D*_Σ_(*t*) could be calculated by the numerical summation of the model’s variables from which the model’s fatality rate was estimated (see Suppl. Mat. 6 and Fig. S9). This rate progressively converges to 1.4% which is poorly consistent with the observations which values range from 4% to 5% (Table S2). Canonical models **K_1_** and **K_2_** directly obtained for the cumulated counts (*C*_Σ_ (*t*) and *D*_Σ_(*t*), respectively) and **K_3_** for their coupling were found much more efficient to reproduce the fatality rate, in particular the canonical model **K_3_** (5.0%) (see Fig. S8). Considering the ability of the virus to propagate easily and silently in both Asia and Europe, and now everywhere in the world and at all society levels, a tremendous number of people will surely be infected by the SARS-Cov-2 in the weeks and months to come, requiring specific measures to slowdown the propagation of the disease. Though, it may probably be extremely difficult to control the disease completely and resurgences must be expected [24] as also suggested by several simulations Fig. 1 and S3.

To analyse the recent outbreaks and compare them to more advanced ones, the global modelling technique was used in its canonical form (Eq. 2) to obtain polynomial models for eight of the mostly affected provinces in China (the Hubei, Zhejiang, Henan, Anhui, Hunan, Jianxi, Guangdong and Heilongjiang) as well as for Japan and South Korea and more recently for Italy. To make the comparison possible between different sizes of population, before modelling, the time series were normalised for a population size of 50 millions inhabitants. Examples of both the original and model phase portraits are shown in Fig. S10 (see also Suppl. Mat. 7). The original and modelled phase portraits of these models are very consistent. These models were obtained at the end of the outbreak for the Chinese provinces and while the epidemic curve had started to decrease in Japan and South Korea; it is why all these models converge to fixed points: their regimes relate to dynamics that are not, -- or not anymore --, chaotic. These were used to perform scenarios for the outbreaks in progress in Europe, in the United States and even more so. On 22 February 2020, several models of canonical form could be also obtained for Italy (see Suppl. Mat. 8) providing another scenario potentially more representative of the European situation.

These models were then used to perform scenarios for fifteen countries as follows. For each country an ensemble of initial states (*C*_Σ_, *C*_1_, *C*_2_)_0_ was estimated from the observational data normalised for 50 million population (initial states were estimated four to eight days before the end of the time series). These initial conditions were then used to perform an ensemble of five simulations for each model. Models incompatible with the present initial conditions were automatically rejected (diverging models). The non rejected simulations are then plotted for comparison with the data observed during the days remaining after the initial conditions. The models the closest to the observations are identified as the most reliable scenarios.

Simulations are provided in Figure three for fifteen countries (the epidemic curves are also provided in Fig. S11), classified from the harder to the smoother situation. These show that many countries have now very largely exceeded the Hubei scenario, in particular Italy for which a model could be obtained much later providing us with another – much harder – scenario. The Italian scenario was even overtaken by Spain and Belgium, and the United Kingdom is now on this scenario. France, the Netherlands and Sweden remain under it, although relatively close to it, Switzerland is significantly lower. The United States appear presently relatively far from the Italian scenario. However, they are still at the beginning of the outbreak in the USA and the growth of infected cases has been particularly quick to overtake the Hubei scenario. Important evolutions still have to be expected there. Among the other countries, Iran and Denmark are significantly over the Hubei scenario whereas Germany, Austria and Norway remain under but close to it. Japan has recently shown a clear restart and is now overtaking the South Korean scenario. Finally, although South Korea could not stop the propagation of the disease, it was able to maintain it at a relatively low level (see the epidemic curves Fig. S11), overtaking its own scenario very slowly.

As already noticed with Japan and as expected for the United States, the present scenarios can evolve. Based on the bulletins [23] published online from 9 February to 2 April 2020, the progressions of these analyses are presented in Figure four. For most European countries, the scenarios have quickly evolved from Heilongjiang to Hubei scenario types and worse ones, highlighting that the outbreaks were not under control in Europe and that strong measures were required to slow down the disease.

## Discussion

The analyses of the control measures taken by the sixteen countries considered in the study show a clear relation between the epidemic level at which the measures were taken (in terms of number of cumulative cases 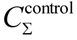 and deaths 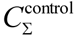, see Table 1 and Fig. S12) and the hardness of the scenarios reached by the countries (as ordered in Figure three). It is still too early to identify the best-case scenario but obviously the earlier the reaction the most likely countries remain on the path of a less dramatic scenario.

Some behaviours require specific comments. In Iran, despite its classification as a moderate scenario, levels 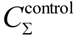 and 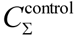 at which control measures were applied are found far much larger than all the other countries. These inconsistencies are difficult to explain since information about the outbreak are scarce and poorly transparent for this country; these high levels probably result from a high under-estimation of the cases of infection and deaths. Despite early control measures taken in Belgium, the spread of the epidemic has significantly overtaken the Italian scenario in terms of cases. This may very likely result from a poor respect of the control measures by one part of the population. Finally, the large dispersion of the control measures in the United States mostly reveals the geographic differences in tackling the outbreak with, in particular, late measures in New York where the epidemic started relatively early, and early measures taken in California, where the epidemic began comparatively later. Since, the epidemic is still at its beginning, the situation will very likely evolve towards harder scenarios in the days to come.

A more in-depth analysis of these results is required to better understand the impact of the type of enforcement (state control *vs* volunteering basis) on the disease evolution outcome, including sociological analysis of the socio-cultural factors influencing control measure implementation between countries (Fig. S13). At this stage, it seems that measures based exclusively on volunteering basis have been efficient to contain the epidemic mostly in South Korea but insufficient to avoid restarts in Japan, and completely ineffective in the Netherlands and the United Kingdom. It shows also that early state controls cannot be sufficient if the measures are not respected by one part of the population.

During the last two weeks, in Europe, the most relevant scenarios have quickly evolved, starting from relatively light situations and evolving to harder and harder scenarios later confirmed step by step (see Fig. 4). Following this path, Italy has largely exceeded the harder situation met in the Hubei province, leading to another, much harder scenario. Spain as well as Belgium have also reached and then exceeded this Italian scenario. They are closely followed by France and the Netherlands. The United Kingdom has also already reached it, and may possibly overtake it in the days to come.

**Fig 4.**
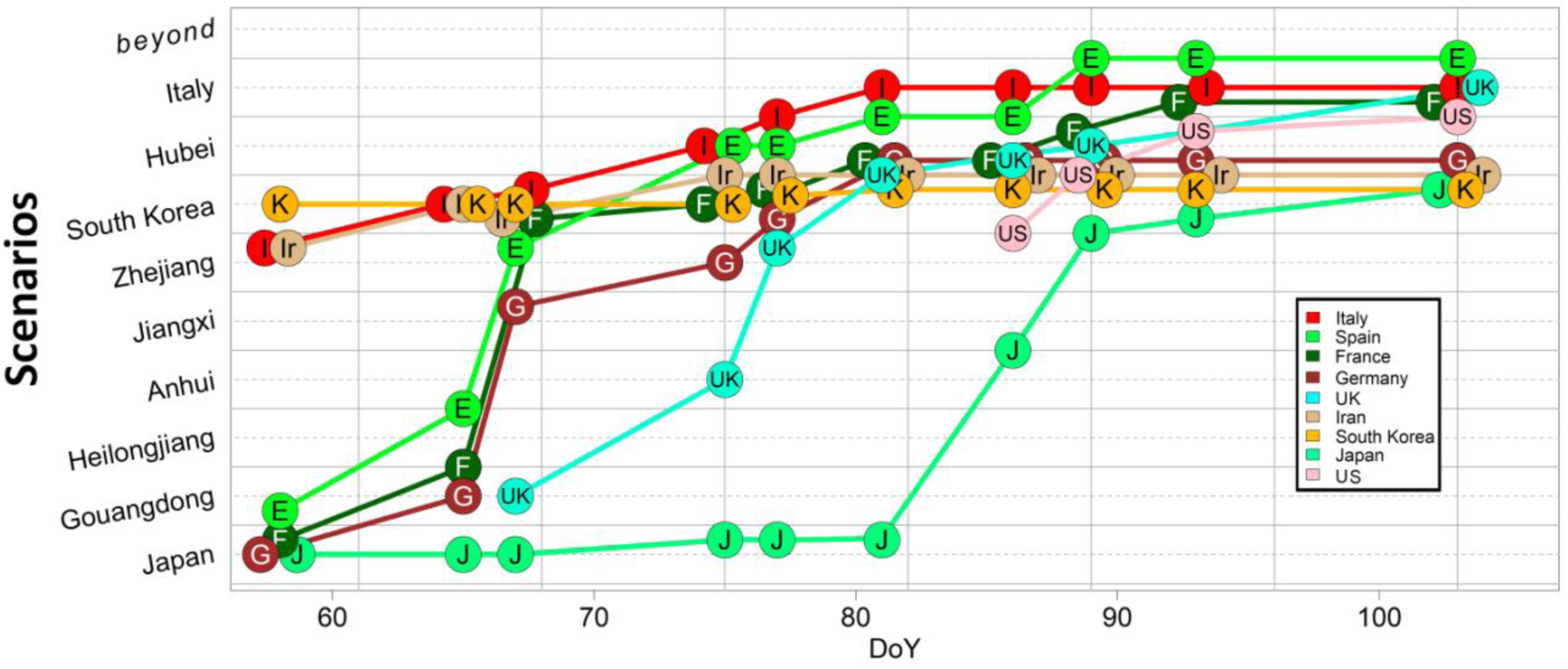
Closest scenarios. Closest scenarios as a function of time for eight countries: South Korea (K), Italy (I), Iran (Ir), Spain (E), France (F), Germany (G), Japan (J), United Kingdom (UK) and the United States of America (US). Results show that the situations can evolve very quickly for the countries who did not take stringent measures to wipe out the epidemic.

One important question that arises from the present results is the scale of applicability. Because the model here obtained at China’s scale is mostly based on a contribution at smaller scale (the Hubei, and more especially Wuhan), it was found to be relatively well applicable to Italy (even without population renormalisation, see Fig.1). The daily deaths toll simulated by the model being even largely exceeded – and more and more – by the Italian observations, it shows that this model is characteristic of an even smaller – intra-province – scale. Indeed, most of the cases and deaths of the Hubei province actually come from the Wuhan district (on 16 March 2020, 73.7% and 79.7%, respectively [24]). As the epidemic could be circumscribed geographically by stringent – and generalised – measures in Wuhan, the model obtained for China finally appears more representative of the extremely smaller urban / suburban scale (~11 millions of inhabitants), a scale comparable to regions such as Lombardy in Italy (~10 M), or cities such as New York City (~5.80 M) and Los Angeles (~10.M) in the United States. Of course, the model cannot exclusively rely on a geographic scale, but also on the conditions in which it was obtained with early and stringent control. Such a hard scenario did happen at a suburb scale under stringent control. Without such a control, the scenarios can get much worse, even at this scale. Therefore, stringent measures similar to those implemented in China but exclusively focused on specific targets may not prevent the development of Hubei scenario types elsewhere. Without control, several scenarios of this type can happen at intra-province scale.

The scenarios here obtained are empirical scenarios. They compare the present epidemic situation to the situations met elsewhere without accounting explicitly for the measures taken to counteract the propagation of the disease. In this sense, the forecasts provided by these scenarios can only be valid provided dynamically equivalent measures are taken (since different measures may have similar effects on the epidemiologic curve by affecting the reproduction number by the same factor). What was observed in practice for the fifteen countries of this study is a quick evolution of all the European scenarios from relatively light (Heilongjiang to Zhejiang) to moderate (South Korea scenario) and then to relatively hard (Hubei) situations and even much harder (the Italian scenario). We have thus reached a situation in Europe with multiple hard Hubei type scenarios at country scale and potentially much beyond in most of the cases. Since the dynamic observed in China is mainly representative of the suburban scale as explained before, several countries did experience multiple hard Wuhan scenarios inside its borders (such as in Italy, Spain or France). The evolution during the last weeks has shown that the United States is following the same evolution from lighter to harder scenarios.

The models obtained for Italy enabled to estimate coming decreasing stages of the epidemic in this country (Fig. S14). It is estimated that a situation with less than 100 new cases per day could be reached by May (see Table S5). Nonetheless, to drop down below this threshold will very likely be quite challenging. Indeed, even in South Korea, where measures have been quicker and more effective, a threshold lower than 50 new cases per day could not be reached. Reaching this stage, specific measures will then be required before getting out from confinement to avoid new clusters restarts.

Nevertheless, the European path may not be the worst possible scenario. The comparison of various outbreaks at a suburban scale (1–10 million inhabitants) can be extremely different from one location to another revealing that, under softer confinement, the situations observed in Italy, Spain or France would have been even worse. Indeed, the evolution observed at four cities of the New York State (New York City, Nassau, Suffolk and Westchester) where the lockdown was late and relatively light, is clearly much worse than what is observed elsewhere, for example in three other cities on the West Coast (King, Santa Clara and Los Angeles), or in four regions of North of Italy (Lombardy, Veneto, Emilia-Romagna and Piedmont), in Daegu metropolitan city in South Korea and in Wuhan (see Fig. S15). These behaviours clearly illustrate that, at such a scale, a late confinement may lead in the United States to scenarios much harder than what was observed in Europe.

The present analyses show that the global modelling approach, possibly in conjunction with other approaches, could be useful for decision makers to monitor the efficiency of control measures and to foresee the extent of the outbreak at various scales. In particular, it could be used to adapt more classical modelling approaches when needed to ensure mitigation and, hopefully, eradication of the disease [5,24] (important methodological differences between the present study and more classical modelling approaches are sketched in Suppl. Mat. 9). This work could be used also to inform decision makers in countries in other parts of the world, especially in the LMICs countries, such as in Africa [25] and South-East Asia, where numbers of Covid-19 cases are still relatively low and where rapid enforcement of control measures should be applied to prevent a catastrophic evolution of the disease.

## Data Availability

The data used in the present study come from the National Health Commission of the Peoples Republic of China and the Johns Hudson University and can be downloaded directly and freely from their respective websites. The GPoM-epidemiologic bulletins were also used. The GPoM tools for global modelling developed in CESBIO by the authors and used to perform the analysis are made available freely and directly downloadable on the Comprehensive R Archrive Network.

http://www.nhc.gov.cn/yjb/pzhgli/new_list.shtml

https://github.com/CSSEGISandData/COVID-19/tree/master/csse_covid_19_data

https://labo.obs-mip.fr/multitemp/bulletin-gpom-epidemiologic/

https://CRAN.R-project.org/package=GPoM

## Acknowledgments

None.

## Author contributions

S.M., M.P. and F.R. designed research; S.M., Y.Z. and M.P. performed research; S.M. and M.H. developed the tools; S.M. and M.P. analysed data; and S.M., M.P., Y.Z., F.R., and Y.K. wrote and edited the paper.

## List of Supplementary Materials

Supplementary Material 1-Detailed contextualization
Supplementary Material 2-Materials and Methods
Supplementary Material 3-Data correction for country inter-comparison
Supplementary Material 4-Dynamical regime
Supplementary Material 5-Other Models **M_2_**, **M_3_**, **K_1_**, **K_2_** and **K_3_**
Supplementary Material 6-Case fatality rate
Supplementary Material 7-Canonical models
Supplementary Material 8-Italy Covid-19 models **I_1_** to **I_5_**
Supplementary Material 9-Comparative analysis

Figs. S1-S15

Tables. S1-S5

